## Supplementary material for "Pre-COVID-19 pandemic health-related behaviours in children (2018-2020) and association with being tested for SARS-CoV-2 and testing positive for SARS-CoV-2 (2020-2021): a retrospective cohort study using survey data linked with routine health data in Wales, UK": Online supplemental appendix 1

Online supplemental appendix 1: see [24]

| Date | Event |
| --- | --- |
| 1 March 2020 | Start of study period. |
| 20 March 2020 | All schools across Wales closed, with the exception of provision of vulnerable children and children of critical workers. |
| 28 April 2020 | More drive-through testing centres. |
| 13 May 2020 | Test, Trace, Protect published including testing strategy to expand from testing workers in hospitals and care comes to symptomatic community testing. |
| 18 May 2020 | Home testing rolled out, enabling symptomatic people to request home coronavirus test. |
| 1 June 2020 | Contract tracing began in Wales. Anyone who tested positive for coronavirus contacted by contact tracer and asked to provide details of everyone they had been in close contact with. Those close contacts contacted and asked to self-isolate for 14 days. |
| 29 June 2020 | Schools opened to pupils from all year groups for limited periods during the week, with only a third of pupils in school at any one time. |
| 15 July 2020 | New Wales coronavirus testing strategy released. |
| 17–24 July 2020 | All schools in Wales closed for the summer holidays. |
| 18 August 2020 | Further investment in testing. Welsh Government announced £32 million funding to improve coronavirus testing including speed of processing tests and ensuring robust testing and contract tracing system for anticipated second wave. |
| 1 September 2020 | Some schools operated a phased return with flexibility to priority groups. |
| 14 September 2020 | Schools opened to all pupils in Wales. |
| 17 September 2020 | Testing update. The Health Minister provided an update on Wales' response to current challenges with coronavirus testing, including a significant increase in demand for testing. |
| 24 September 2020 | NHS COVID-19 app launched for people aged 16 and over, including increased capacity for identifying contacts of those tested positive for coronavirus. |
| 29 September 2020 | Wales' Minister for Health and Social Care sets out the prioritisation for Covid-19 testing in Wales as the Welsh Government move into a new phase of its response. The Minster set out six priorities for testing, with those working in education or childcare settings the fifth priority group and all symptomatic individuals being sixth. |
| 19–30 October 2020 | Autumn half term holiday (three local authorities had 2 week half term) |
| 2 November 2020 | Pupils in year 9 and above not expected to be present in school due to firebreak in Wales. |
| 16 November 2020 | New financial support scheme launched for people who need to self-isolate due to a positive coronavirus test result or those asked to do so by NHS Wales Test, Trace, Protect. |
| 18 November 2020 | Merthyr Tydfil (one of 22 local authorities) to be first whole area testing pilot in Wales. Everyone offered Covid-19 testing, whether symptomatic or asymptomatic. The mass testing programme used LFTs. |
| 27 November 2020 | Mass testing extended (use of LFTs). |
| 14 December 2020 | All secondary schools in Wales moved to online remove learning for last week of term before Christmas (14-18 December 2020). Many primary schools also closed. |
| 14 December 2020 | Plans for serial testing in schools. The Welsh Government announces plans to roll out serial testing in schools and colleges from January 2021. |
| 18 December 2020 | Testing infrastructure developments to support mass testing of symptomatic people across the Welsh population. In addition to increasing testing infrastructure for people with symptoms, also developing approach and support for people without symptoms to access lateral flow tests (LFT). |

Online supplemental appendix 1: see [24]

| | |
|---|---|
| 4 January 2021 | All schools across Wales closed and moved to online remote learning, with the exception of provision for vulnerable children and children of critical workers. |
| 28 January 2021 | Publication of an updated coronavirus testing strategy including to continue to test symptomatic individuals. |
| 1 February 2021 | Users of NHS Covid-19 app could apply for self-isolation payment. People on low incomes who are asked to self-isolate via the NHS Covid-19 are eligible to apply for the £500 self-isolation support payment, alongside those who have been asked to self-isolate by the Test, Trace, Protect service and parents/carers whose child has been asked to self-isolate by their education setting. |
| 5 February 2021 | Twice weekly testing in schools. The Health Minister announces that daily contact testing in schools and colleges will be paused and instead, twice weekly testing using LFTs. Guidance for detection of positive LFT encourages follow up PCR test. |
| 27 February 2021 | Testing offer extended to upper secondary and college learners. Welsh Government Ministers announce that they are extending the offer of regular, twice weekly, LFTs at home to all those of upper secondary age. This will start with offering tests to years 11 to 13, and to all further education college learners and those on work-based apprenticeship and traineeship programmes. |
| 10 March 2021 | Testing for close contacts of positive cases. Wales' Health Minister, announces that people who are close contacts of someone who has tested positive for coronavirus and have been asked to isolate by contact tracers will now be offered a coronavirus test. Also extra £50 million to allow health boards to extend contact tracing over the summer. |
| 15 March 2021 | All remaining primary school pupils and secondary pupils in qualification years 11 and 13 able to return to learning on site. Other secondary years were able to return for check ins. |
| 15 March 2021 | Voluntary asymptomatic testing offer available to secondary age pupils in years 10 and above. |
| 22 March 2021 | Community asymptomatic testing programme extended to end of September 2021. |
| 30 March 2021 | Updated testing strategy published. |
| 12 April 2021 | All remaining pupils were able to return to learning on site. |
| 13 April 2021 | Voluntary asymptomatic testing offer extended to all secondary school age years (years 7 and above). |
| 26 May 2021 | Covid tests (LFTs) encouraged for people holidaying in Wales. |
| 31 August 2021 | End of study period. |
