## Supplementary material for "Pre-COVID-19 pandemic health-related behaviours in children (2018-2020) and association with being tested for SARS-CoV-2 and testing positive for SARS-CoV-2 (2020-2021): a retrospective cohort study using survey data linked with routine health data in Wales, UK": Online supplemental appendix 3

### THE HAPPEN SURVEY

* Required

Consent Form

Before you start please click this link to read the information sheet -> https://happen-wales.co.uk/wp-content/uploads/2019/02/Child-Consent-2019.pdf

1. I have read the child information sheet -> https://happen-wales.co.uk/wp-content/uploads/2019/02/Child-Consent-2019.pdf (click the link if you haven't read it) and understand that if I take part I can change my mind at any time, and this will not be a problem at all. *

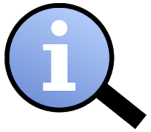

*Mark only one oval.*

- ◯ Yes
- ◯ No

2. I am happy for you to use my questionnaire for research. Only the researchers in the team will know my name and will not tell anyone else my answers *

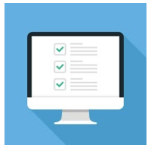

*Mark only one oval.*

- ◯ Yes
- ◯ No do not use my questionnaire

3. I am happy for you to look at my school and health records to see how my school is doing (as a group). This is anonymous which means I cannot be identified *

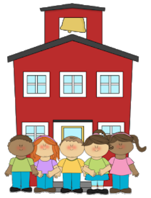

*Mark only one oval.*

- ◯ Yes
- ◯ No

If you do not wish to take part in the questionnaire please do not continue.

Please click next to start the questionnaire!

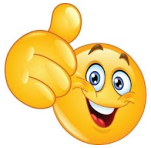

#### About You

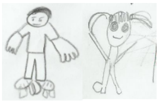

4. First Name *

____________________

5. Last Name *

____________________

6. Home Post Code *

____________________

7. What school do you go to? *

____________________

8. What year are you in? *

*Mark only one oval.*

- ◯ Year 4
- ◯ Year 5
- ◯ Year 6
- ◯ Year 7

9. Gender *

*Mark only one oval.*

- ◯ Boy
- ◯ Girl
- ◯ Prefer not to say

Date of Birth

10. Year *

*Mark only one oval.*

- ◯ 2007
- ◯ 2008
- ◯ 2009
- ◯ 2010
- ◯ 2011
- ◯ 2012

11. Month *

*Mark only one oval.*

- ( ) January
- ( ) February
- ( ) March
- ( ) April
- ( ) May
- ( ) June
- ( ) July
- ( ) August
- ( ) September
- ( ) October
- ( ) November
- ( ) December

12. Day *

*Mark only one oval.*

- ( ) 1
- ( ) 2
- ( ) 3
- ( ) 4
- ( ) 5
- ( ) 6
- ( ) 7
- ( ) 8
- ( ) 9
- ( ) 10
- ( ) 11
- ( ) 12
- ( ) 13
- ( ) 14
- ( ) 15
- ( ) 16
- ( ) 17
- ( ) 18
- ( ) 19
- ( ) 20
- ( ) 21
- ( ) 22
- ( ) 23
- ( ) 24
- ( ) 25
- ( ) 26
- ( ) 27
- ( ) 28
- ( ) 29
- ( ) 30
- ( ) 31

#### YESTERDAY

CHAT
Child Health and Activity Tool

Firstly, think carefully about what you did YESTERDAY and then answer the following questions....

13. 1. What did you eat for breakfast YESTERDAY? *

*Check all that apply.*

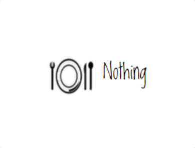


- [ ] Nothing

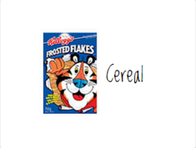


- [ ] Sugary cereal e.g. cocopops, frosties, sugar puffs, chocolate cereals

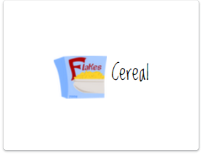


- [ ] Healthy cereal e.g. porridge, weetabix, readybrek, muesli, branflakes, cornflakes

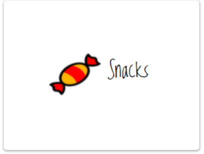


- [ ] Snacks

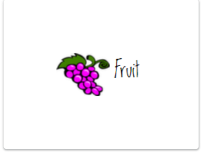


- [ ] Fruit

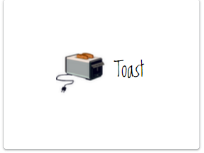


- [ ] Toast

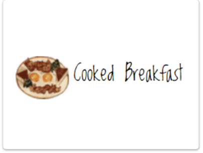


- [ ] Cooked breakfast

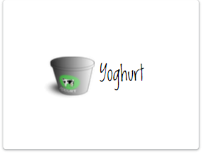


- [ ] Yoghurt

Other: [ ] ____________

14. 2. How did you get to school YESTERDAY morning? *

*Mark only one oval.*

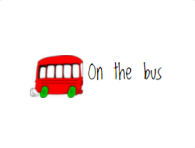


( ) On the bus

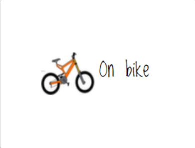


( ) On bike

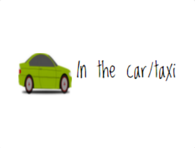


( ) In the car/taxi

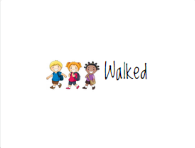


( ) Walked

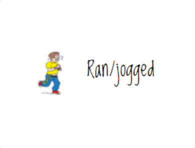


( ) Ran/jogged

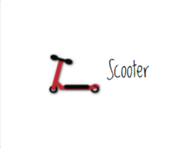


( ) Scooter

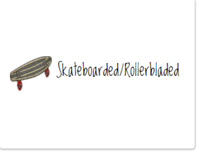


( ) Skateboarded/Rollerbladed

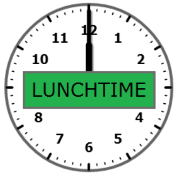


15. 3. What did you have to eat for lunch YESTERDAY? *

*Mark only one oval.*

( ) School dinner
( ) Packed lunch
( ) Nothing

16. 4. What did you do for MOST of your break-times YESTERDAY? (This includes lunchtime) *

*Mark only one oval.*

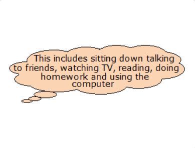


( ) Sat around inside or outside

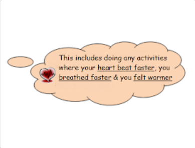


( ) Ran around

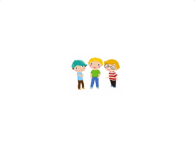

( ) Stood around

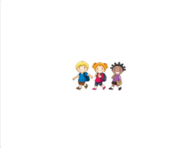

( ) Walked around

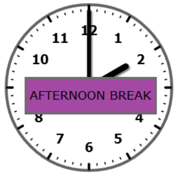


17. 5. Do you have an afternoon break at school? *

*Mark only one oval.*

( ) YES
( ) NO

18. 6. How did you get home YESTERDAY? *

*Mark only one oval.*

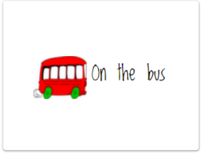


- On the bus

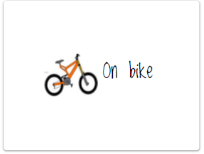


- On bike

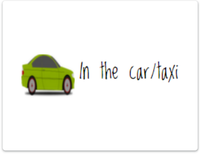


- In the car/taxi

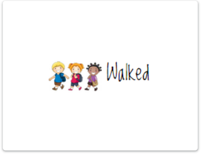


- Walked

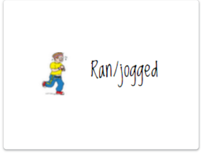


- Ran/jogged

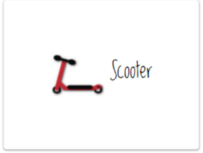


- Scooter

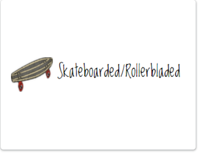


- Skateboarded/Rollerbladed

AFTER SCHOOL

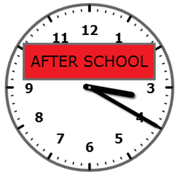


19. 7. How many portions of fruit and vegetables did you eat YESTERDAY? *

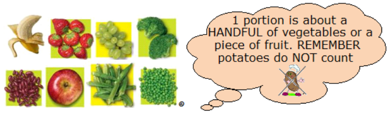


*Mark only one oval.*

- 0
- 1
- 2
- 3
- 4
- 5
- 6
- 7
- 8

20. 8. How many times did you brush your teeth YESTERDAY? *

*Mark only one oval.*

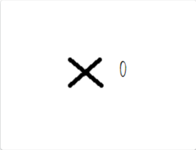


- 0

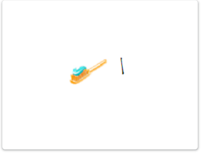


- 1

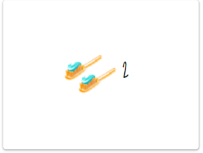


- 2

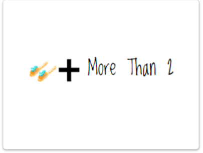


- 3

21. 9. What time did you fall asleep YESTERDAY (to the nearest half hour)? *

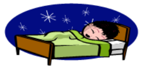

*Mark only one oval.*

- 7:00pm
- 7:30pm
- 8:00pm
- 8:30pm
- 9:00pm
- 9:30pm
- 10:00pm
- 10:30pm
- 11:00pm
- 11:30pm
- 12:00am
- 12:30am
- 1:00am
- 1:30am
- 2:00am
- 3:00am
- 3:30am
- 4:00am

22. 10. What time did you wake up TODAY (to the nearest half hour)? *

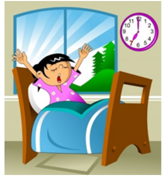

*Mark only one oval.*

- ◯ 5:00am
- ◯ 5:30am
- ◯ 6:00am
- ◯ 6:30am
- ◯ 7:00am
- ◯ 7:30am
- ◯ 8:00am
- ◯ 8:30am
- ◯ 9:00am

#### THE LAST WEEK

NOW think about what you did in the last 7 days...

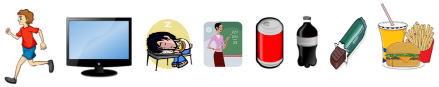

23. 11a. In the last 7 days, how many days did you do sports or exercise for at least 1 hour in total (This includes doing any activities or playing sports where your heart beat faster, you breathed faster and you felt warmer? *

*Mark only one oval.*

- ◯ 0 days
- ◯ 1-2 days
- ◯ 3-4 days
- ◯ 5-6 days
- ◯ 7 days

24. 11b. In the last 7 days, how many days did you watch TV/play online games/use the internet etc. for 2 or more hours a day (in total)? *

*Mark only one oval.*

- ◯ 0 days
- ◯ 1-2 days
- ◯ 3-4 days
- ◯ 5-6 days
- ◯ 7 days

25. 11c. In the last 7 days, how many days did you feel tired? *

*Mark only one oval.*

- ◯ 0 days
- ◯ 1-2 days
- ◯ 3-4 days
- ◯ 5-6 days
- ◯ 7 days

26. 11d. In the last 7 days, how many days did you feel like you could concentrate/pay attention well in class? *

*Mark only one oval.*

- ◯ 0 days
- ◯ 1-2 days
- ◯ 3-4 days
- ◯ 5-6 days
- ◯ 7 days

27. 11e. In the last 7 days, how many days did you drink at least one fizzy drink (e.g. coke, fanta, sprite) *

*Mark only one oval.*

- ◯ 0 days
- ◯ 1-2 days
- ◯ 3-4 days
- ◯ 5-6 days
- ◯ 7 days

28. 11f. In the last 7 days, how many days did you eat at least one sugary snack (e.g. chocolate bar, sweets) *

*Mark only one oval.*

- ◯ 0 days
- ◯ 1-2 days
- ◯ 3-4 days
- ◯ 5-6 days
- ◯ 7 days

29. 11g. In the last 7 days, how many days did you eat take away foods (e.g. McDonalds, KFC, chinese) *

*Mark only one oval.*

- ◯ 0 days
- ◯ 1-2 days
- ◯ 3-4 days
- ◯ 5-6 days
- ◯ 7 days

#### Sport and Activity

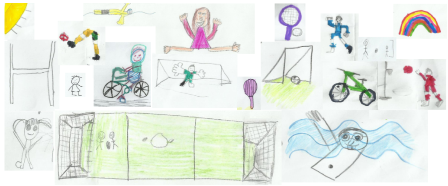

30. 12. These questions are going to ask you how you feel about physical activity (This includes any activity where your heart beats faster, you breathe faster and you feel warmer) *

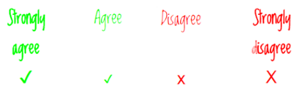


*Mark only one oval per row.*

| | Strongly agree | Agree | Disagree | Strongly disagree |
|---|---|---|---|---|
| I want to take part in physical activity | ◯ | ◯ | ◯ | ◯ |
| I feel confident to take part in lots of different physical activities | ◯ | ◯ | ◯ | ◯ |
| I am good at lots of different physical activities | ◯ | ◯ | ◯ | ◯ |
| I understand why taking part in physical activity is good for me | ◯ | ◯ | ◯ | ◯ |

31. 13a. How many times do you take part in a sports club OUTSIDE OF SCHOOL each week?

*Mark only one oval.*

| 0 | 1 | 2 | 3 | 4 | 5 | 6 | 7 | 8 | 9 | 10 |
|---|---|---|---|---|---|---|---|---|---|---|
| ◯ | ◯ | ◯ | ◯ | ◯ | ◯ | ◯ | ◯ | ◯ | ◯ | ◯ |

32. 13b. If you take part in a sports club OUTSIDE of school, what is the name of the sports club? (For example Swansea Rugby Club Under 11's)

_______________________________

33. 14. Are you a member of cubs, brownies, scouts or guides? *

*Mark only one oval.*

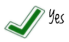


◯ Yes

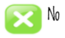


◯ No

34. 15. Which of these sports or physical activities would you MOST like to try? (That you haven't tried before) *

*Mark only one oval.*

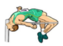

◯ Athletics

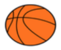

◯ Basketball

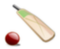

◯ Cricket

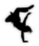

◯ Dance

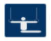

◯ Gymnastics

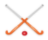

◯ Hockey

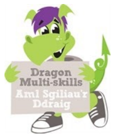


◯ Multi Skills

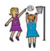

◯ Netball

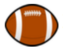

◯ Rugby

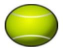

◯ Tennis

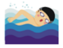

◯ Swimming

◯ I do not want to try anything-I don't like sport or activity

◯ Other: _______________

35. 16. Can you ride a bike WITHOUT STABILISERS? *

*Mark only one oval.*

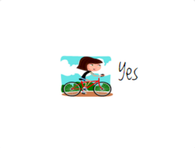


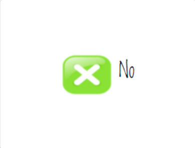


( ) Yes
( ) No

36. 17. Can you swim 25 metres WITHOUT A FLOAT OR ARMBANDS? (This is 1 length of a standard swimming pool) *

*Mark only one oval.*

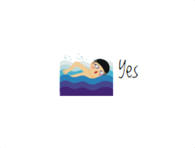


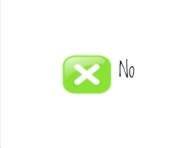


( ) Yes
( ) No

#### You and your feelings

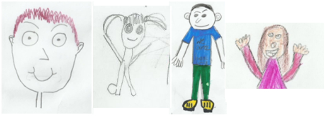

This part of the survey is going to ask you how you feel. There are no right or wrong answers. You should just pick the answer which is best for you.

37. 18. Tell us if you agree or disagree with the following: *

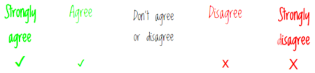


*Mark only one oval per row.*

| | Strongly agree | Agree | Don't agree or disagree | Disagree | Strongly disagree |
|---|---|---|---|---|---|
| I am doing well at school | ( ) | ( ) | ( ) | ( ) | ( ) |
| I have lots of choice over things that are important to me | ( ) | ( ) | ( ) | ( ) | ( ) |
| There are lots of things I'm good at | ( ) | ( ) | ( ) | ( ) | ( ) |

19. On a scale of 0 to 10 (0 being very unhappy and 10 being very happy, how do you feel about:

*Based on the Good Childhood Index by the Children's Society

38. Your Health *

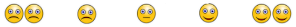

*Mark only one oval.*

| | 0 | 1 | 2 | 3 | 4 | 5 | 6 | 7 | 8 | 9 | 10 | |
|---|---|---|---|---|---|---|---|---|---|---|---|---|
| Very unhappy | ( ) | ( ) | ( ) | ( ) | ( ) | ( ) | ( ) | ( ) | ( ) | ( ) | ( ) | Very happy |

39. Your School *

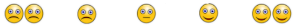

*Mark only one oval.*

| | 0 | 1 | 2 | 3 | 4 | 5 | 6 | 7 | 8 | 9 | 10 | |
|---|---|---|---|---|---|---|---|---|---|---|---|---|
| Very unhappy | ( ) | ( ) | ( ) | ( ) | ( ) | ( ) | ( ) | ( ) | ( ) | ( ) | ( ) | Very happy |

40. Your Family *

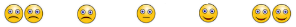

*Mark only one oval.*

| | 0 | 1 | 2 | 3 | 4 | 5 | 6 | 7 | 8 | 9 | 10 | |
|---|---|---|---|---|---|---|---|---|---|---|---|---|
| Very unhappy | ( ) | ( ) | ( ) | ( ) | ( ) | ( ) | ( ) | ( ) | ( ) | ( ) | ( ) | Very happy |

41. Your Friends *

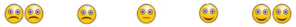

*Mark only one oval.*

| | 0 | 1 | 2 | 3 | 4 | 5 | 6 | 7 | 8 | 9 | 10 | |
|---|---|---|---|---|---|---|---|---|---|---|---|---|
| Very unhappy | ( ) | ( ) | ( ) | ( ) | ( ) | ( ) | ( ) | ( ) | ( ) | ( ) | ( ) | Very happy |

42. Your Appearance (how you look) *

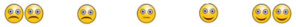

*Mark only one oval.*

| | 0 | 1 | 2 | 3 | 4 | 5 | 6 | 7 | 8 | 9 | 10 | |
|---|---|---|---|---|---|---|---|---|---|---|---|---|
| Very unhappy | ( ) | ( ) | ( ) | ( ) | ( ) | ( ) | ( ) | ( ) | ( ) | ( ) | ( ) | Very happy |

43. Your Life *

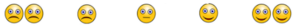

*Mark only one oval.*

| | 0 | 1 | 2 | 3 | 4 | 5 | 6 | 7 | 8 | 9 | 10 | |
|---|---|---|---|---|---|---|---|---|---|---|---|---|
| Very unhappy | ( ) | ( ) | ( ) | ( ) | ( ) | ( ) | ( ) | ( ) | ( ) | ( ) | ( ) | Very happy |

#### You and your Feelings

Based on the Me and My Feelings Questionnaire ( Deighton, Tymms, Vostanis, Belsky, Fonagy, Brown, Martin, Patalay, & Wolpert, 2012)

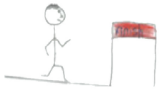

44. 20. Remember, there are no right or wrong answers, just pick which is right for you. *

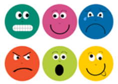

*Mark only one oval per row.*

| | Never | Sometimes | Always |
|---|---|---|---|
| I feel lonely | ◯ | ◯ | ◯ |
| I cry a lot | ◯ | ◯ | ◯ |
| I am unhappy | ◯ | ◯ | ◯ |
| I feel nobody likes me | ◯ | ◯ | ◯ |
| I worry a lot | ◯ | ◯ | ◯ |
| I have problems sleeping | ◯ | ◯ | ◯ |
| I wake up in the night | ◯ | ◯ | ◯ |
| I am shy | ◯ | ◯ | ◯ |
| I feel scared | ◯ | ◯ | ◯ |
| I worry when I am at school | ◯ | ◯ | ◯ |
| I get very angry | ◯ | ◯ | ◯ |
| I lose my temper | ◯ | ◯ | ◯ |
| I hit out when I am angry | ◯ | ◯ | ◯ |
| I do things to hurt people | ◯ | ◯ | ◯ |
| I am calm | ◯ | ◯ | ◯ |
| I break things on purpose | ◯ | ◯ | ◯ |

#### Your Local Area

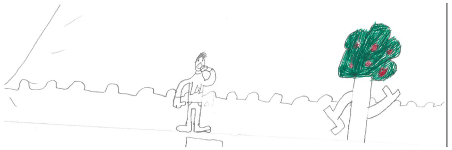

45. 21. On a scale of 0 to 10 (0 being not very safe and 10 being very safe), how safe do you feel playing in your area? *

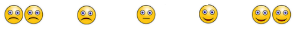

*Mark only one oval.*

| | 0 | 1 | 2 | 3 | 4 | 5 | 6 | 7 | 8 | 9 | 10 | |
|---|---|---|---|---|---|---|---|---|---|---|---|---|
| Not very safe | ◯ | ◯ | ◯ | ◯ | ◯ | ◯ | ◯ | ◯ | ◯ | ◯ | ◯ | Very safe |

46. 22a. From your house, can you walk to school?

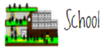


*Mark only one oval.*

◯ Yes
◯ No

47. 22b. From your house, can you easily walk to a park?

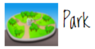


*Mark only one oval.*

◯ Yes
◯ No

48. 22c. From your house, can you easily walk to a leisure centre/sports centre?

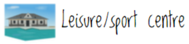


*Mark only one oval.*

◯ Yes
◯ No

49. 23. Are you happy with the area that you live in?

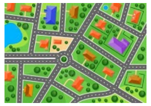

*Mark only one oval.*

◯ Yes
◯ No

24. If you could change something to make you and your friends healthier and happier, what would you change...

50. IN SCHOOL? *

_______________________________________________
_______________________________________________
_______________________________________________
_______________________________________________
_______________________________________________

51. OUT OF SCHOOL? *

_______________________________________________
_______________________________________________
_______________________________________________
_______________________________________________
_______________________________________________

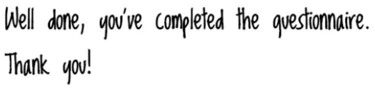


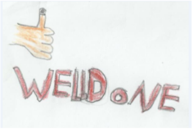


Don't forget to press submit below!

This content is neither created nor endorsed by Google.

Google Forms
