## Supplementary material for "Pre-COVID-19 pandemic health-related behaviours in children (2018-2020) and association with being tested for SARS-CoV-2 and testing positive for SARS-CoV-2 (2020-2021): a retrospective cohort study using survey data linked with routine health data in Wales, UK": Online supplemental appendix 4

**Online supplemental appendix 4: HAPPEN survey variable codebook**

| Exposures | HAPPEN Survey item | Responses | Analyses coding |
|---|---|---|---|
| **Ate breakfast** | *13. What did you eat for breakfast yesterday?* | *Nothing*<br>*Cereal*<br>*Snacks*<br>*Fruit*<br>*Toast*<br>*Cooked breakfast*<br>*Yoghurt* | Binary:<br>*1 = Cereal; Snacks; Fruit; Toast; Cooked breakfast; Yoghurt*<br>*0 = Nothing* |
| **Active travel to school** | *14. How did you get to school yesterday morning?* | *On the bus*<br>*In the car/taxi*<br>*Walked*<br>*On bike*<br>*Ran/jogged*<br>*Scooter*<br>*Skateboarded/rollerbladed* | Binary:<br>*1 = Walked; On bike; Ran/jogged; Scooter; Skateboarded/rollerbladed*<br>*0 = On the bus; In the car/taxi* |
| **Active travel from school** | *18. How did you get home yesterday?* | *On the bus*<br>*In the car/taxi*<br>*Walked*<br>*On bike*<br>*Ran/jogged*<br>*Scooter*<br>*Skateboarded/rollerbladed* | Binary:<br>*1 = Walked; On bike; Ran/jogged; Scooter; Skateboarded/rollerbladed*<br>*0 = On the bus; In the car/taxi* |
| **Toothbrush 2+ per day** | *20. How many times did you brush your teeth yesterday?* | *0 – 3* | Continuous:<br>*0 – 3* |
| **5+ fruit and veg** | *19. How many portions of fruit and vegetables did you eat yesterday?* | *0 – 8* | Continuous:<br>*0 – 8* |

**Online supplemental appendix 4: HAPPEN survey variable codebook**

| | | | |
|---|---|---|---|
| **Sleep 9+ hours** | *21. What time did you fall asleep last night* | *(30 min intervals)*<br>*7:00pm – 4:00am* | Continuous:<br>*Sleep hours calculated from item 21 and 22* |
| | *22. What time did you wake up this morning?* | *(30 min intervals)*<br>*5:00am – 9:00am* | |
| **Physically active 60+ mins every day previous 7 days** | *23. In the last 7 days, how many days did you do sports or exercise for at least 1 hour in total (This includes doing any activities or playing sports where your heart beat faster, you breathed faster and you felt warmer* | *0 days*<br>*1 – 2 days*<br>*3 – 4 days*<br>*5 – 6 days*<br>*7 days* | Ordinal:<br>*0 days*<br>*1 – 2 days*<br>*3 – 4 days*<br>*5 – 6 days*<br>*7 days* |
| **Sedentary/screen time 2 hours every day previous 7 days** | *24. In the last 7 days, how many days did you watch TV/play online games/use the internet etc. for 2 or more hours a day (in total)?* | *0 days*<br>*1 – 2 days*<br>*3 – 4 days*<br>*5 – 6 days*<br>*7 days* | Ordinal:<br>*0 days*<br>*1 – 2 days*<br>*3 – 4 days*<br>*5 – 6 days*<br>*7 days* |
| **Tired 7 days** | *25. In the last 7 days, how many days did you feel tired?* | *0 days*<br>*1 – 2 days*<br>*3 – 4 days*<br>*5 – 6 days*<br>*7 days* | Ordinal:<br>*0 days*<br>*1 – 2 days*<br>*3 – 4 days*<br>*5 – 6 days*<br>*7 days* |

**Online supplemental appendix 4: HAPPEN survey variable codebook**

| | | | |
|---|---|---|---|
| **Sugary snack 7 days** | *28. In the last 7 days, how many days did you eat at least one sugary snack (e.g. chocolate bar, sweets)* | *0 days*<br>*1 – 2 days*<br>*3 – 4 days*<br>*5 – 6 days*<br>*7 days* | Ordinal:<br>*0 days*<br>*1 – 2 days*<br>*3 – 4 days*<br>*5 – 6 days*<br>*7 days* |
| ***Participate in at least 3 out of school clubs*** | *31. How many times do you take part in a sports club OUTSIDE OF SCHOOL each week?* | *0 - 10* | Continuous:<br>*0 - 10* |
| ***Can ride a bike*** | *35. Can you ride a bike without stabilisers?* | *No*<br>*Yes* | Binary:<br>*1 = Yes*<br>*0 = No* |
| ***Can swim 25m*** | *36. Can you swim 25 metres without a float or armbands (This is 1 length of a standard swimming pool)* | *No*<br>*Yes* | Binary:<br>*1 = Yes*<br>*0 = No* |
| ***Age on 01/03/2020*** | *Decimal age on 1 March 2020* | Continuous | Continuous |
| ***Sex*** | *Sex* | *Girl*<br>*Boy* | Binary:<br>*0 = Girl*<br>*1 = Boy* |
| ***WIMD*** | *Welsh Index of Multiple Deprivation 2019* | | Coding framework from WIMD 2019[34] |
