## Supplementary material for "Pre-COVID-19 pandemic health-related behaviours in children (2018-2020) and association with being tested for SARS-CoV-2 and testing positive for SARS-CoV-2 (2020-2021): a retrospective cohort study using survey data linked with routine health data in Wales, UK": Online supplemental appendix 5

**Online supplemental appendix 5:** Full descriptive statistics table by tested for SARS-CoV-2 and tested positive for SARS-CoV-2.

| | | **Tested for SARS-CoV-2 n (%)** | **Not tested for SARS-CoV-2 n (%)** | **Tested positive for SARS-CoV-2 n (%)** | **No evidence of positive SARS-CoV-2 test n (%)** |
|---|---|---|---|---|---|
| **Sample** | | 39.1% (2,764) | 60.9% (4,298) | 8.1% (569) | 91.9% (6,498) |
| **Age at time of HAPPEN survey** | | 10.1 ± 0.8 | 9.9 ± 0.9 | 10.1 ± 0.8 | 9.9 ± 0.8 |
| **Age on 01/03/2020 (start of period of interest)** | | 10.6 ± 0.9 | 10.3 ± 1.1 | 10.6 ± 1.0 | 10.4 ± 1.0 |
| **Sex** | **Boy** | 49.3% (1,363) | 46.7% (2,005) | 44.3% (252) | 48.0% (3,116) |
| | **Girl** | 48.9% (1,352) | 51.8% (2,226) | 54.5% (310) | 50.3% (3,268) |
| | *Missing* | *1.8% (49)* | *1.5% (67)* | 1.2% (7) | 1.7% (109) |
| **WIMD 2019 quintiles** | **1 (most deprived)** | 24.3% (672) | 23.9% (1,025) | 28.5% (162) | 23.6% (1,535) |
| | **2** | 19.9% (551) | 19.02% (826) | 19.7% (112) | 19.5% (1,265) |
| | **3** | 16.5% (455) | 17.4% (748) | 17.6% (100) | 17.0% (1,103) |
| | **4** | 15.6% (431) | 15.8% (678) | 14.1% (80) | 15.9% (1,029) |
| | **5 (least deprived)** | 18.0% (497) | 16.8% (771) | 16.5% (94) | 17.3% (1,124) |
| | *Missing* | *5.7% (158)* | *7.0% (300)* | *3.7% (21)* | *6.7% (5437)* |
| | | **Previous day** | | | |
| **Ate breakfast** | **Yes** | 93.0% (2,571) | 92.1% (3,797) | 93.4% (538) | 92% (6,012) |
| | **No** | 7% (193) | 7.3% (319) | 5.6% (31) | 7.3% (481) |
| | *Missing* | *0%* | *0%* | *0%* | *0%* |
| **Active travel to school** | **Yes** | 38.5% (1,065) | 39.8% (1,710) | 37.6% (214) | 39.4% (2,561) |
| | **No** | 61.5% (1,699) | 60.2% (2,588) | 62.4% (355) | 60.6% (3,932) |

| | | | | | |
|---|---|---|---|---|---|
| | *Missing* | *0%* | *0%* | *0%* | *0%* |
| **Active travel from school** | **Yes** | 43.0% (1,187) | 43.0% (1,846) | 42.4% (241) | 43.0% (2,792) |
| | **No** | 57.0% (1,577) | 57.0% (2,452) | 57.6% (328) | 57.0% (3,701) |
| | *Missing* | *0%* | *0%* | *0%* | *0%* |
| **Toothbrush continuous** | **0** | 3.3% (91) | 3.4% (146) | 1.9% (11) | 3.5% (227) |
| | **1** | 20.0% (552) | 21.0% (903) | 18.6% (106) | 20.6% (1,358) |
| | **2** | 67.1% (1,854) | 65.2% (2,802) | 69.6% (396) | 65.2% (4,294) |
| | **3** | 9.6% (265) | 10.3% (446) | 9.5% (54) | 10.0% (659) |
| | *Missing* | *0.1% (<5)* | *<0.1% (<5)* | *0.4% (<5)* | *<0.1%% (<5)* |
| **Fruit/veg portions (continuous)** | **0** | 14.3% (395) | 15.3% (657) | 12.5% (71) | 15.1% (981) |
| | **1** | 16.1% (445) | 17.4% (749) | 15.8% (90) | 17.0% (1,104) |
| | **2** | 17.7% (489) | 17.5% (754) | 19.5% (111) | 17.4% (1,132) |
| | **3** | 17.5% (484) | 16.5% (711) | 16.7% (95) | 16.9% (1,110) |
| | **4** | 12.7% (351) | 11.9% (510) | 13.5% (77) | 12.1% (784) |
| | **5** | 10.5% (291) | 10.6% (455) | 11.8% (67) | 10.4% (679) |
| | **6** | 4.5% (123) | 4.3% (186) | 2.8% (16) | 4.5% (293) |
| | **7** | 2.3% (63) | 2.1% (92) | 4.2% (24) | 2.0% (131) |
| | **8** | 4.5% (123) | 4.3% (184) | 3.2% (18) | 4.5% (289) |
| | *Missing* | 0% | 0% | *0%* | *0%* |
| **Sleep hours** | | 9.4 ± 1.6 | 9.4 ± 1.6 | 9.4 ± 1.6 | 9.4 ± 1.6 |
| **Number of days physically active ≥ 60 minutes** | **0** | 6.5% (179) | 7.9% (339) | 4.0% (23) | 7.6% (495) |
| | **1-2 days** | 27.9% (772) | 29.0% (1,246) | 27.8% (158) | 28.7% (1,860) |
| | **3-4 days** | 27.5% (761) | 26.2% (1,128) | 30.9% (176) | 26.4% (1,712) |
| | **5-6 days** | 18.3% (505) | 17.0% (731) | 18.1% (103) | 17.5% (1,133) |

| | | | | | |
|---|---|---|---|---|---|
| | **7 days** | 19.8% (557) | 19.9% (854) | 19.2% (109) | 19.9% (1,292) |
| | *Missing* | *0%* | *0%* | *0%* | *0%* |
| **Number of days sedentary/screen time ≥ two hours** | **0** | 5.2% (144) | 6.1% (262) | 5.5% (31) | 5.8% (375) |
| | **1-2 days** | 24.2% (674) | 23.5% (1,011) | 24.8% (141) | 23.8% (1,544) |
| | **3-4 days** | 21.7% (599) | 20.6% (886) | 21.1% (120) | 21.0% (1,365) |
| | **5-6 days** | 14.0% (386) | 13.8% (593) | 13.9% (79) | 13.9% (900) |
| | **7 days** | 34.8% (961) | 36.0% (1,546) | 34.8% (198) | 35.6% (2,309) |
| | *Missing* | *0%* | *0%* | *0%* | *0%* |
| **Number of days tired** | **0** | 21.0% (582) | 21.0% (903) | 19.2% (109) | 21.2% (1,376) |
| | **1-2 days** | 32.4% (895) | 32.0% (1,377) | 35.7% (203) | 31.9% (2,069) |
| | **3-4 days** | 17.6% (487) | 17.5% (754) | 18.8% (107) | 17.5% (1,134) |
| | **5-6 days** | 10.0% (276) | 9.3% (399) | 10.5% (60) | 9.5% (615) |
| | **7 days** | 19.0% (524) | 20.1% (865) | 15.8% (90) | 20.0% (1,299) |
| | *Missing* | *0%* | *0%* | *0%* | *0%* |
| **Number of days sugary snack** | **0** | 6.5% (179) | 7.7% (332) | 6.3% (36) | 7.3% (475) |
| | **1-2 days** | 34.9% (964) | 32.7% (1,407) | 35.0% (199) | 33.5% (2,172) |
| | **3-4 days** | 25.3% (698) | 26.7% (1,146) | 25.1% (143) | 26.2% (1,701) |
| | **5-6 days** | 13.4% (371) | 12.0% (515) | 15.3% (87) | 12.3% (799) |
| | **7 days** | 20.0% (552) | 20.9% (898) | 18.3% (104) | 20.7% (1,346) |
| | *Missing* | *0%* | *0%* | *0%* | *0%* |
| | | | **General** | | |
| **Number of out of school clubs** | **0** | 27.7% (766) | 32.3% (1,387) | 25.1% (143) | 31.0% (2,010) |
| | **1** | 17.9% (495) | 16.9% (726) | 16.0% (91) | 17.4% (1,130) |
| | **2** | 16.0% (443) | 15.1% (650) | 14.9% (85) | 15.5% (1,008) |
| | **3** | 11.1% (308) | 10.4% (446) | 13.3% (76) | 10.4% (678) |

| | | | | | |
|---|---|---|---|---|---|
| | **4** | 7.4% (204) | 7.3% (313) | 7.6% (43) | 7.3% (474) |
| | **5** | 6.2% (171) | 5.8% (251) | 5.8% (33) | 6.0% (389) |
| | **6** | 3.4% (95) | 2.5% (109) | 5.1% (29) | 2.7% (175) |
| | **7** | 3.3% (91) | 2.5% (107) | 5.1% (29) | 2.6% (169) |
| | **8** | 1.1% (29) | 0.8% (33) | 1.8% (10) | 0.8% (52) |
| | **9** | 0.9% (24) | 0.7% (32) | 1.2% (7) | 0.8% (49) |
| | **10** | 3.9% (107) | 4.0% (174) | 3.3% (19) | 4.0% (262) |
| | *Missing* | *1.1% (31)* | *1.6% (70)* | *0.7% (<5)* | *1.5% (97)* |
| **Can ride a bike** | **Yes** | 88.8% (2,444) | 86.0% (3,696) | 91.4% (520) | 86.7% (5,641) |
| | **No** | 11.2% (309) | 14.0% (602) | 8.6% (49) | 13.3% (862) |
| | *Missing* | *0%* | *0%* | *0%* | *0%* |
| **Can swim 25m** | **Yes** | 78.9% (2,180) | 72.9% (3,134) | 80.3% (457) | 74.8% (4,857) |
| | **No** | 21.1% (584) | 27.1% (1,164) | 19.7% (112) | 25.2% (1,636) |
| | *Missing* | *0%* | *0%* | *0%* | *0%* |
