## Supplementary material for "Pre-COVID-19 pandemic health-related behaviours in children (2018-2020) and association with being tested for SARS-CoV-2 and testing positive for SARS-CoV-2 (2020-2021): a retrospective cohort study using survey data linked with routine health data in Wales, UK": Online supplemental appendix 6

**Online supplemental appendix 6:**
Multivariable logistic regression model of health behaviour markers and probability of PCR-test without confounders.

| **PCR tested for SARS-CoV-2 (n=6,958, $R^2$=0.01)** | **OR** | ***p* value** | **95% CI** |
|---|---|---|---|
| **Ate breakfast** | 1.05 | 0.632 | 0.87 – 1.27 |
| *Reference: did not eat breakfast* | 1.00 | | |
| **Active travel to school** | 0.92 | 0.238 | 0.80 – 1.06 |
| *Reference: did not active travel to school* | 1.00 | | |
| **Active travel from school** | 1.08 | 0.273 | 0.94 – 1.24 |
| *Reference: did not active travel from school* | 1.00 | | |
| **Number of fruit/vegetable portions** | 1.00 | 0.941 | 0.98 – 1.03 |
| *Reference: 0 fruit/vegetable portions* | 1.00 | | |
| **Number of times teeth brushed** | 0.97 | 0.474 | 0.90 – 1.05 |
| *Reference: did not brush teeth* | 1.00 | | |
| **Sleep hours** | 0.99 | 0.654 | 0.96 – 1.02 |
| *Reference: 0 days physically active ≥60 mins (previous seven days)* | 1.00 | | |
| **1-2 days physically active ≥ 60 mins** | 1.12 | 0.276 | 0.91 – 1.38 |
| **3-4 days physically active ≥ 60 mins** | 1.14 | 0.221 | 0.92 – 1.42 |
| **5-6 days physically active ≥ 60 mins** | 1.17 | 0.177 | 0.93 – 1.47 |
| **7 days physically active ≥ 60 mins** | 1.09 | 0.475 | 0.87 – 1.37 |
| *Reference: 0 days sedentary ≥ two hours (previous seven days)* | 1.00 | | |
| **1-2 days sedentary ≥ two hours** | 1.16 | 0.209 | 0.92 – 1.46 |
| **3-4 days sedentary ≥ two hours** | 1.18 | 0.166 | 0.93 – 1.49 |
| **5-6 days sedentary ≥ two hours** | 1.15 | 0.275 | 0.90 – 1.47 |
| **7 days sedentary ≥ two hours** | 1.14 | 0.256 | 0.91 – 1.44 |
| *Reference: 0 days felt tired (previous seven days)* | 1.00 | | |
| **1-2 days felt tired** | 0.98 | 0.791 | 0.86 – 1.13 |
| **3-4 days felt tired** | 0.99 | 0.881 | 0.84 – 1.16 |
| **5-6 days felt tired** | 1.04 | 0.667 | 0.86 – 1.26 |
| **7 days felt tired** | 0.97 | 0.730 | 0.83 – 1.14 |
| *Reference: 0 days consumed sugary snack (previous seven days)* | | | |
| **1-2 days consumed sugary snack** | 1.21* | 0.062 | 0.99 – 1.49 |
| **3-4 days consumed sugary snack** | 1.08 | 0.489 | 0.87 – 1.33 |
| **5-6 days consumed sugary snack** | 1.29** | 0.034 | 1.02 – 1.63 |
| **7 days consumed sugary snack** | 1.12 | 0.314 | 0.90 – 1.39 |
| **Number of out of school clubs participation** | 1.02 | 0.121 | 1.00 to 1.04 |
| **Can ride a bike** | 1.16* | 0.064 | 0.99 – 1.35 |

| | | | |
|---|---|---|---|
| *Reference: cannot ride a bike* | 1.00 | | |
| **Can swim 25m** | 1.30** | 0.000 | 1.15 – 1.46 |
| *Reference: cannot swim 25m* | 1.00 | | |

OR: Odds Ratio; 95% CI: 95% confidence intervals; p<0.05**, p<0.1*. See online supplemental appendix 4 for variable codebook.

Multivariable logistic regression model of health behaviour markers and probability of PCR-test positive without confounders.

| PCR test positive for SARS-CoV-2 (n=6,958, $R^2$=0.01) | OR | *p* value | 95% CI |
|---|---|---|---|
| **Ate breakfast** | 1.30 | 0.170 | 0.89 – 1.91 |
| *Reference: did not eat breakfast* | 1.00 | | |
| **Active travel to school** | 0.91 | 0.451 | 0.71 – 1.17 |
| *Reference: did not active travel to school* | 1.00 | | |
| **Active travel from school** | 1.07 | 0.614 | 0.83 – 1.36 |
| *Reference: did not active travel from school* | 1.00 | | |
| **Number of fruit/vegetable portions** | 0.99 | 0.574 | 0.94 – 1.03 |
| *Reference: 0 fruit/vegetable portions* | 1.00 | | |
| **Number of times teeth brushed** | 1.07 | 0.385 | 0.92 – 1.24 |
| *Reference: did not brush teeth* | 1.00 | | |
| **Sleep hours** | 0.97 | 0.266 | 0.92 – 1.02 |
| *Reference: 0 days physically active ≥ 60 mins (previous seven days)* | 1.00 | | |
| **1-2 days physically active ≥ 60 mins** | 1.71 | 0.023 | 1.08 – 2.73 |
| **3-4 days physically active ≥ 60 mins** | 1.87 | 0.009 | 1.17 – 2.99 |
| **5-6 days physically active ≥ 60 mins** | 1.61 | 0.059 | 0.98 – 2.63 |
| **7 days physically active ≥ 60 mins** | 1.49 | 0.117 | 0.91 – 2.43 |
| *Reference: 0 days sedentary ≥ two hours (previous seven days)* | 1.00 | | |
| **1-2 days sedentary ≥ two hours** | 1.03 | 0.877 | 0.68 – 1.57 |
| **3-4 days sedentary ≥ two hours** | 1.00 | 0.983 | 0.66 – 1.54 |
| **5-6 days sedentary ≥ two hours** | 1.01 | 0.958 | 0.65 – 1.59 |
| **7 days sedentary ≥ two hours** | 1.10 | 0.660 | 0.72 – 1.66 |
| *Reference: 0 days felt tired (previous seven days)* | 1.00 | | |
| **1-2 days felt tired** | 1.21 | 0.125 | 0.95 – 1.55 |
| **3-4 days felt tired** | 1.17 | 0.278 | 0.88 – 1.55 |
| **5-6 days felt tired** | 1.21 | 0.273 | 0.86 – 1.69 |
| **7 days felt tired** | 0.92 | 0.600 | 0.69 – 1.24 |
| *Reference: 0 days consumed sugary snack (previous seven days)* | 1.00 | | |
| **1-2 days consumed sugary snack** | 1.14 | 0.499 | 0.78 – 1.67 |
| **3-4 days consumed sugary snack** | 1.03 | 0.873 | 0.70 – 1.53 |
| **5-6 days consumed sugary snack** | 1.38 | 0.131 | 0.91 – 2.11 |
| **7 days consumed sugary snack** | 1.04 | 0.867 | 0.69 – 1.56 |
| **Number of out of school clubs participation** | 1.05 | 0.007 | 1.01 – 1.09 |
| **Can ride a bike** | 1.40 | 0.032 | 1.03 – 1.92 |

| | | | |
|---|---|---|---|
| *Reference: cannot ride a bike* | 1.00 | | |
| **Can swim 25m** | 1.16 | 0.207 | 0.92 – 1.45 |
| *Reference: cannot swim 25m* | 1.00 | | |
